# Genomic SEM Applied to Explore Etiological Divergences in Bipolar Subtypes

**DOI:** 10.1101/2023.04.29.23289281

**Authors:** Jeremy M. Lawrence, Sophie Breunig, Isabelle F. Foote, Connor B. Tallis, Andrew D. Grotzinger

**Affiliations:** Institute for Behavioral Genetics, University of Colorado Boulder, Boulder, CO; Department of Psychology and Neuroscience, University of Colorado Boulder, Boulder, CO

## Abstract

**Background:** Bipolar Disorder (BD) is an overarching diagnostic class defined by the presence of at least one prior manic episode (BD I) or both a prior hypomanic episode and a prior depressive episode (BD II). Traditionally, BD II has been conceptualized as a less severe presentation of BD I, however, extant literature to investigate this claim has been mixed.

**Methods:** We apply Genomic Structural Equation Modeling (Genomic SEM) to investigate divergent genetic pathways across BD’s two major subtypes using the most recent GWAS summary statistics from the PGC. We begin by identifying divergences in genetic correlations across 89 external traits using a Bonferroni corrected threshold. We also use a theoretically informed follow-up model to examine the extent to which the genetic variance in each subtype is explained by schizophrenia and major depression. Lastly, Transcriptome-wide SEM (T-SEM) was used to identify gene expression patterns associated with the BD subtypes.

**Results:** BD II was characterized by significantly larger genetic overlap with internalizing traits (e.g., neuroticism, insomnia, physical inactivity), while significantly stronger associations for BD I were limited. Consistent with these findings, the follow-up model revealed a much larger major depression component for BD II. T-SEM results revealed 41 unique genes associated with risk pathways across BD subtypes.

**Conclusions:** Divergent patterns of genetic relationships across external traits provide support for the distinction of the bipolar subtypes. However, our results also challenge the illness severity conceptualization of BD given stronger genetic overlap across BD II and a range of clinically relevant traits and disorders.

## Introduction

Bipolar disorder is an overarching diagnostic class with two primary subtypes: bipolar I disorder (BD I) and bipolar II disorder (BD II). BD I is characterized by at least one prior manic episode and BD II by both a prior hypomanic and major depressive episode (American Psychiatric Association, 2013). The operationalization of these subtypes reflects conceptualizations of BD I and II as part of an illness continuum ranging from schizoaffective to unipolar depression, with BD I lying closer to the former and BD II to the latter (Gershon, 1982). Although BD I is traditionally considered the more clinically severe version of these two disorders, extant research to support this claim, or more generally distinguish these two subtypes beyond their diagnostic definitions, is limited and mixed. For example, comparisons of BD I and BD II have revealed mixed findings regarding whether these subtypes display differences in comorbidity patterns (Baek et al., 2011; Loftus et al., 2020), suicidal behaviors (Dunner, 2004; Karanti et al., 2020; Novick et al., 2010), or neuroimaging outcomes (Hozer & Houenou, 2016). In addition, while the broader bipolar diagnostic class is associated with various negative health outcomes, including physical inactivity and related comorbidities (McElroy & Keck, 2012), it is unclear whether these associations are primarily driven by one of the two subtypes. The current study investigates the etiological differences that characterize the underlying genetic risk pathways of these two major BD subtypes.

Challenging the model that BD I is simply a more severe version of BD II, genetic epidemiology has found that the subtypes “breed true.” That is, relatives of individuals with a particular BD subtype exhibit heightened risk for that subtype, indicating risk pathways are at least partially unique to a given BD subtype (Andreasen, 1987; Heun & Maier, 1993). In the past decade, genome-wide association studies (GWAS) of complex phenotypes have started to unpack the specific genetic variants associated with different outcomes. The most recent bipolar GWAS from the Psychiatric Genomics Consortium (PGC) uncovered 64 significantly associated genetic variants (Mullins et al., 2021). In addition, GWAS data for each BD subtype was used to examine the overall genetic signal for BD I and BD II, revealing SNP-based heritability estimates 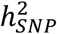 of 21.2% and 11.6%, respectively, and a genetic correlation (*r*_*g*_) of 0.85. This *r*_*g*_ was significantly different from 1, indicating the presence of subtype-specific signal. However, research into what might explain this divergent genetic signal is limited.

Bivariate genomic methods such as LD-score regression (LDSC; Bulik-Sullivan et al., 2015) allow for estimating genetic overlap across two traits with GWAS summary statistics. As these GWAS summary statistics are often publicly available and need not come from the same participant sample, this offers the unique opportunity to investigate genetic overlap across a wide range of rare and even mutually exclusive outcomes. Indeed, progress in understanding BD subtypes using phenotypic approaches has likely been stymied, at least in part, by the pragmatic difficulty of recruiting a participant sample of sufficient size for both subtypes. Genomic Structural Equation Modeling (Genomic SEM; Grotzinger et al., 2019) is a multivariate framework for modeling the genetic overlap estimated from LDSC. In the present study, we apply Genomic SEM and its recent extensions to examine genetic convergence and divergence across bipolar subtypes. At the genome-wide level, we explicitly model and statistically compare the genetic correlations across BD subtypes and a range of cognitive, health, interpersonal, and psychiatric outcomes. We go on to apply Transcriptome-wide SEM (Grotzinger et al., 2022) to identify genes whose expression is associated with general or subtype-specific BD risk. These results provide biological support for many of the clinically observed differences in subtype-specific outcome measures, aid in the identification of elevated risk factors associated with each subtype, and further our understanding of the etiological convergences and divergences across bipolar subtypes.

## Methods

### GWAS Datasets

GWAS summary statistics for BD I (25,060 cases, 449,978 controls) and II (6,781 cases, 364,075 controls) were utilized from the most recent PGC Freeze 3 GWAS (Mullins et al., 2021). Cases were defined using either medical records or internationally recognized diagnostic manuals (i.e., DSM-IV, ICD-9, ICD-10) administered by trained interviewers and clinicians. We utilize publicly available European ancestry summary statistics for 95 external traits, reflecting a collection of outcomes deemed to be psychiatrically relevant across the broad categories of cognitive, interpersonal, and social functioning, physical health, and psychiatric outcomes. Among this initial pool of 95 external traits, 89 had 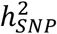 *Z*-statistics greater than the recommended cut-off of 4 put forth by the original LDSC developers for producing interpretable estimates of genetic covariance (Bulik-Sullivan et al., 2015). These 89 traits were carried forward for Genomic SEM analyses. A summary of each dataset used in this study with sample characteristics (i.e., case/control numbers) is provided in Supplementary Table 1.

### Genomic SEM

A standard set of quality control (QC) filters was first applied to all GWAS summary statistics using the *munge* function within Genomic SEM. This included filtering to HapMap3 SNPs and, when this information was available, removing SNPs with a minor allele frequency (MAF) < .01 and with an imputation score (INFO) < 0.9. These “munged” summary statistics were subsequently used as input to the multivariable version of *ldsc* implemented in Genomic SEM, which estimates the genetic covariance and sampling covariance matrices across included traits. The genetic covariance matrix contains SNP-based heritability estimates on the diagonal and genetic covariances on the off-diagonal. For binary traits, these estimates were converted to the more interpretable liability scale using the population prevalence from the corresponding publication and the sum of effective sample size across cohorts contributing to the GWAS (Grotzinger et al., 2023). The sampling covariance contains squared standard errors on the diagonal (the sampling variances) and sampling covariances on the off-diagonal, which index sampling dependencies that will arise when there is participant sample overlap. LD weights used to estimate the regression model in LDSC were obtained from 1000 Genomes Phase 3 European LD Scores, excluding the major histocompatibility complex (MHC) due to complex LD structures in this region that can bias estimates.

Before running models, the genetic covariance and sampling covariance matrices were transformed into genetic correlation and sampling correlation matrices. This was done to examine differences in the proportion of genetic overlap with other traits, accounting for differences in the overall 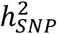 of the two BD subtypes (results examining the genetic covariances are reported in Supplementary Table 2). These standardized LDSC matrices were then used as input to a series of models specified within Genomic SEM. The first was a saturated model in which the genetic overlap across the BD subtypes and each external trait were estimated. Next, a constrained model was specified in which the relationships between each BD subtype and the external trait were fixed to both be equal. The *p*-value associated with the constrained model’s *χ*^2^ statistic indicates the decrement in model fit, relative to the fully saturated model, as a consequence of imposing the equality constraint. Significant values then indicate that the BD subtypes have significantly different genetic correlations (*r*_*g*_) with the external trait. Separate models were run for each external trait and a Bonferroni corrected threshold (*p* < 5.6E-4 = .05/89) was applied to determine statistical significance.

Finally, we fit a series of theoretically informed models with MDD and SCZ as correlated predictors of both BD subtypes in a genetic multiple regression model. We examined four sets of model constraints for this model: *(i)* MDD and SCZ with regression paths to BD I fixed to be equal, *(ii)* MDD and SCZ with regression paths to BD II fixed to be equal, *(iii)* regression paths from MDD to BD I and II fixed to be equal, and *(iv)* regression paths from SCZ to BD I and II fixed to be equal. Model comparisons were not done in sequential order. That is, for each of the four models, all other parameters were freely estimated aside from the two parameters with an imposed equality constraint. A final model was then estimated that included any equality constraints that could be imposed without a significant decrement in model fit. Significance was defined here using a Bonferroni corrected threshold for the four comparisons of *p <* .0125. The genetic variance for each BD subtype for this model was partitioned based on the genetic signal shared with MDD and SCZ. The residual or unique variance of each subtype was then further broken down into shared and unique residual genetic variance between the subtypes.

### Transcriptome-wide SEM

We utilized Transcriptome-wide SEM (T-SEM) to identify tissue-specific gene expression shared and unique to BD I and II (Grotzinger et al., 2022). Univariate transcriptome-wide association studies (TWAS) uncover the relationship between gene-expression and a single trait through summary-based transcriptomic imputation. T-SEM reflects a multivariate extension of TWAS that allows for examining unique and shared gene-expression across constellations of genetically overlapping traits.

First, FUSION was used to perform univariate, summary-based TWAS for both BD subtypes (Gusev et al., 2016). 16 sets of publicly available functional weights were utilized from three data resources: *(i)* the Genotype-Tissue Expression project (GTEx v8), with functional weights for 13 brain tissue types, *(ii)* the CommonMind Consortium (CMC), with functional weights for two dorsolateral prefrontal cortex datasets, and *(iii)* PsychEncode, with functional weights for the prefrontal cortex (Fromer et al., 2016; Gandal et al., 2018; The GTEx Consortium, 2020). In total, this yielded 84,698 genes with imputed expression across tissue types. These univariate FUSION statistics were then used as input to the *read_fusion* function in Genomic SEM, which backs out the gene expression-phenotype covariances that are standardized with respect to the phenotypic variance. This allows these gene expression estimates to be added to the LDSC genetic covariance matrices as they are now on the same scale. Finally, the *userGWAS* function was used to estimate the effect of gene expression on a latent BD factor defined by the two BD subtypes. As this was a two-indicator factor, the factor loadings of the two BD subtypes were constrained to be equal in order to ensure model identification.

These analyses produced two sets of output: (*i*) the relationship between gene expression and the latent BD factor and (*ii*) a Q_Gene_ statistic that identifies patterns of gene expression that do not operate via the factor and for this model are thereby likely to be subtype-specific in their effect. We have previously demonstrated via simulation that Q_Gene_ is typically significant when a gene has directionally discordant associations with the traits that define the factor or shows a much stronger association with one trait (Grotzinger et al., 2022). Hits on the factor were defined as genes that were significant at a Bonferroni corrected threshold of *p <* 5.90E-7 that were not also significant for Q_Gene_. Finally, over-representation analysis (ORA) was conducted using the *WebGestalt* package (Liao et al., 2019) to identify overlapping gene sets between signal enriched for the BD factor and external traits.

## Results

### Genetic Overlap with External Traits

Among the 89 examined external traits, 18 surpassed the Bonferroni corrected significance threshold. Genetic correlations across the BD subtypes and the traits with the top 20 lowest model *χ*^2^ comparison *p*-values (*χ*^2^ *p*) are presented in Figure 1. A full list of genetic correlations is presented in Supplementary Table 3. Across a broad range of psychiatric, interpersonal, and health-related outcomes, significant divergences in the underlying genetic risk pathways between each BD subtype are observed, with BD II consistently exhibiting greater levels of shared risk with external outcomes. We consider more specific results across different domains of external traits directly below.

**Fig. 1.**
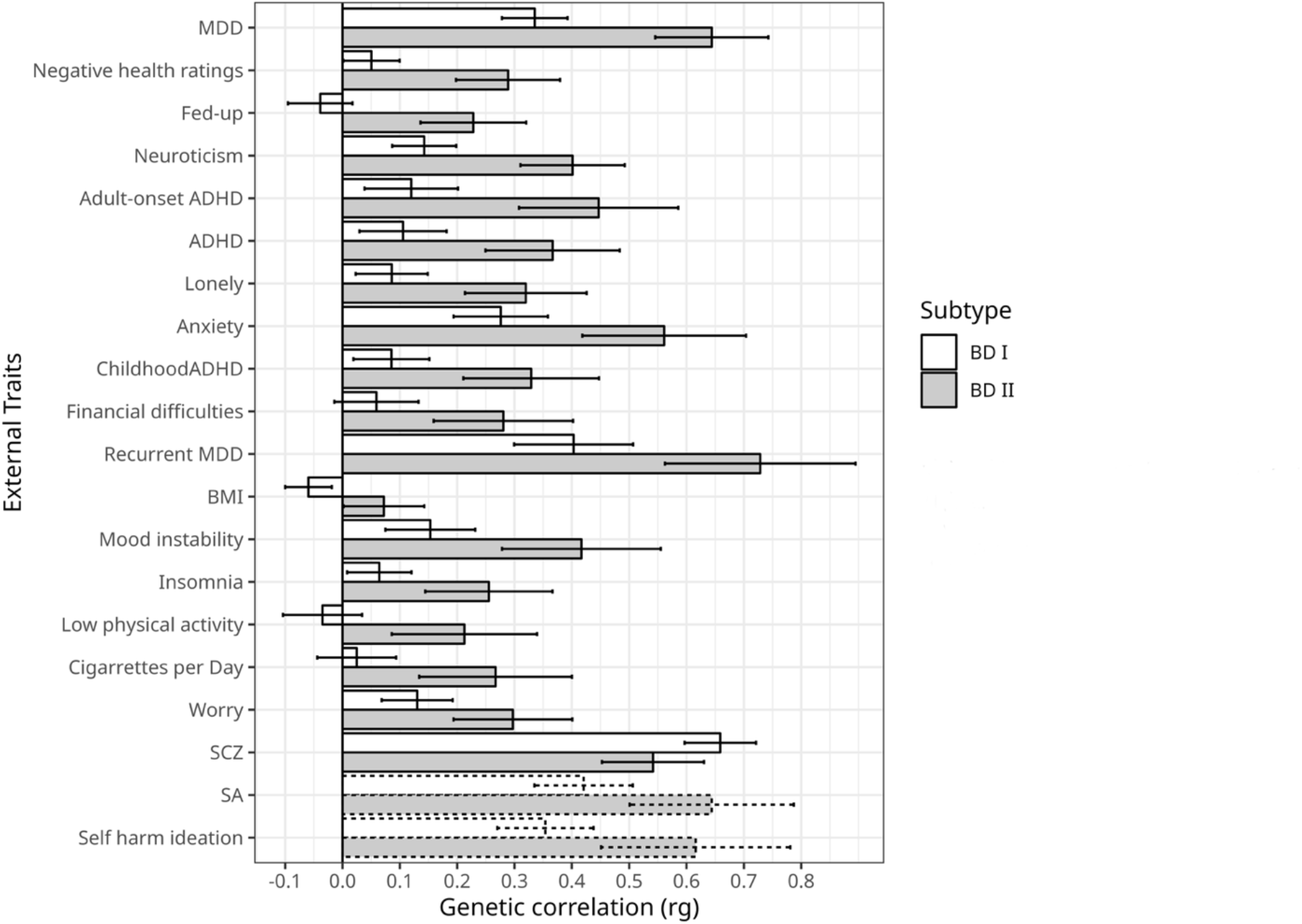
Genetic correlations between BD subtypes and external traits. Traits sorted by ascending *p-*values. Error bars depict 95% confidence intervals. Dashed bars represent traits not surpassing a Bonferroni corrected significance threshold.

### Psychiatric Disorders & Symptoms

The one disorder that showed a stronger relationship with BD I was SCZ (BD I: *r*_*g*_ *=* .66, *SE* = .03; BD II: *r*_*g*_ *=* .54, *SE* = .05, *χ*^2^ *p* = 5.18E-04), though the relationship with BD II can still be described as sizeable. Conversely, we identified several differences where BD II exhibited elevated risk-sharing relative to BD I across a range of disorders. For example, BD II had higher genetic overlap with ADHD (BD I: *r*_*g*_ *=* .11, *SE* = .04; BD II: *r*_*g*_ *=* .37, *SE* = .06, *χ*^2^ *p* = 2.85E-07), a finding that replicated when splitting across ADHD diagnosed in childhood (BD I: *r*_*g*_ *=* .09, *SE* = .03; BD II: *r*_*g*_ *=* .33, *SE* = .06, *χ*^2^ *p* = 7.53E-06), and adulthood (BD I: *r*_*g*_ *=* .12, *SE* = .04; BD II: *r*_*g*_ *=* .45, *SE* = .07, *χ*^2^ *p* = 1.04E-07). BD II also showed a stronger relationship with a number of disorders in the internalizing space, including anxiety disorders (BD I: *r*_*g*_ *=* .28, *SE* = .04; BD II: *r*_*g*_ *=* .56, *SE* = .07, *χ*^2^ *p* = 2.12E-06), MDD (BD I: *r*_*g*_ *=* .34, *SE* = .03; BD II: *r*_*g*_ *=* .64, *SE* = .05, *χ*^2^ *p* = 4.06E-14), and recurrent MDD (BD I: *r*_*g*_ *=* .40, *SE* = .05; BD II: *r*_*g*_ *=* .73, *SE* = .09, *χ*^2^ *p* = 2.37E-05).

In addition to the disorders themselves, several differences were observed between BD subtypes and psychiatric symptoms or closely linked personality traits. These differences can generally be described as reflecting a stronger association between different aspects of internalizing behavior and BD II relative to BD. This included: insomnia (BD I: *r*_*g*_ *=* .06, *SE* = .03; BD II: *r*_*g*_ *=* .26, *SE* = .06, *χ*^2^ *p* = 5.78E-05); feeling fed up (BD I: *r*_*g*_ *=* -.04, *SE* = .03; BD II: *r*_*g*_ *=* .23, *SE* = .05, *χ*^2^ *p* = 1.34E-09); mood instability (BD I: *r*_*g*_ *=* .15, *SE* = .04; BD II: *r*_*g*_ *=* .42, *SE* = .07, *χ*^2^ *p* = 4.48E-05); feelings of worry (BD I: *r*_*g*_ *=* .13, *SE* = .03; BD II: *r*_*g*_ *=* .30, *SE* = .05, *χ*^2^ *p* = 3.46E-04); and neuroticism (BD I: *r*_*g*_ *=* .14, *SE* = .03; BD II: *r*_*g*_ *=* .40, *SE* = .05, *χ*^2^ *p* = 1.47E-09). The point estimates for BD II were also larger for suicide attempts (BD I: *r*_*g*_ *=* .42, *SE* = .04; BD II: *r*_*g*_ *=* .64, *SE* = .07, *χ*^2^ *p* = 6.73E-04) and ever considering self-harm (BD I: *r*_*g*_ *=* .35, *SE* = .04; BD II: *r*_*g*_ *=* .62, *SE* = .08, *χ*^2^ *p* = 1.45E-03), although these estimates did not surpass the Bonferroni corrected significance threshold.

Our theoretically informed follow-up analysis with MDD and SCZ as correlated predictors of each subtype provides further context to this pattern of results. All four equality constraints resulted in significant decrements in model fit *(p*s ≤ .0125), indicating that MDD and SCZ do not exhibit equal genetic overlap within or across the BD subtypes. The fully saturated multiple regression model with no constraints was used to partition the genetic variance of each BD subtype into the following groups: the genetic variance explained by MDD; the genetic variance explained by SCZ; the genetic variance shared between MDD and SCZ; and the genetic variance shared between (i.e., shared residuals) and unique (i.e., unique residuals) to each subtype (Figure 2). The genetic variance in BD I reflected: 37.9% (*SE* = 5.5%) SCZ, 1.7% (*SE* = 1.0%) MDD, 5.3% (*SE* = 1.3%) shared variance between MDD and SCZ, 22.8% (*SE* = 4.7%) shared residual variance with BD II, and 32.3% (*SE* = 4.1%) residual variance unique of BD II. BD II was partitioned into: 13.5% (*SE* = 4.3%) SCZ, 27.2% (*SE* = 6.7%) MDD, 12.8% (*SE* = 1.7%) shared variance between MDD and SCZ, 19.3% (*SE* = 4.9%) shared residual variance with BD I, and 27.3% (*SE* = 4.2%) residual variance unique of BD I.

**Fig. 2.**
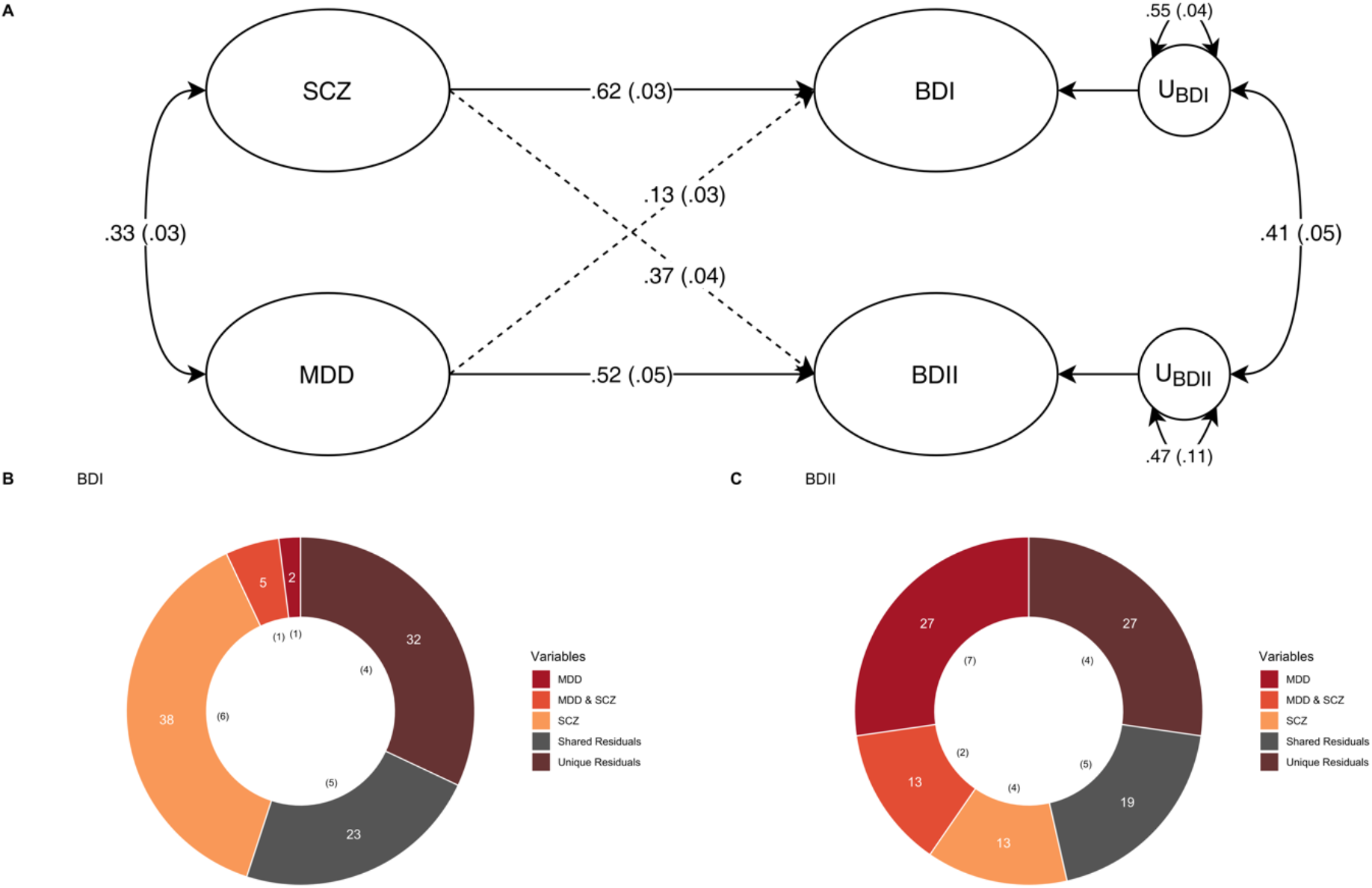
Major Depression and Schizophrenia as Correlated Predictors of the Bipolar Subtypes. **(A)** Standardized results using Genomic SEM to construct a model with MDD and SCZ as correlated predictors of BD I & II. **(B, C)** Percent variance within BD I and II accounted for by signal unique to MDD, unique to SCZ, shared by MDD and SCZ, unique to each subtype (unique residuals) and shared between the subtypes (shared residuals). Numbers in parentheses in both panels reflect the corresponding standard error.

### Interpersonal & Social Functioning

We also observed patterns of increased genetic risk shared between BD II and outcomes related to interpersonal and social functioning. More specifically, BD II was more strongly associated with increased loneliness (BD I: *r*_*g*_ *=* .09, *SE* = .03; BD II: *r*_*g*_ *=* .32, *SE* = .05, *χ*^2^ *p* = 1.18E-06) and financial difficulties (BD I: *r*_*g*_ *=* .06, *SE* = .04; BD II: *r*_*g*_ *=* .28, *SE* = .06, *χ*^2^ *p* = 1.20E-05).

### Physical Health Traits

BD II displayed greater overlap with physical health traits often related to features of major depression. These differences include significantly stronger relationships between BD II and negative self-ratings of health (BD I: *r*_*g*_ *=* .05, *SE* = .03; BD II: *r*_*g*_ *=*.29, *SE* = .05, *χ*^2^ *p* = 1.74E-10), physical inactivity (BD I: *r*_*g*_ *=* -.04, *SE* = .04; BD II: *r*_*g*_ *=* .21, *SE* = .07, *χ*^2^ *p* = 7.44E-05), and number of cigarettes smoked per day (BD I: *r*_*g*_ *=* .03, *SE* = .04; BD II: *r*_*g*_ *=*.27, *SE* = .07, *χ*^2^ *p* = 1.08E-04).

#### T-SEM

For each BD subtype, 84,684 tissue-specific gene expression estimates were obtained. Univariate TWAS for each of the disorders revealed 261 hits for BD I and one hit for BD II. T-SEM revealed 86 hits on the BD factor (Figure 3 for Miami plot; Supplementary Table 4 for list of all T-SEM hits). As many genes are present across multiple tissues, these 86 hits reflect 41 unique gene IDs. The top 5 most significant hits for the BD factor were: *DCLK3, PELI3, LINC02033, GNL3*, and *ZSCAN9*. Follow up ORA analyses indicated significant enrichment for the BD factor gene expression hits and a gene-set previously implicated in the overarching bipolar diagnostic class, *p* = 1.83E-6 (Liao et al., 2019). Supplementary Figure 1 visually depicts that T-SEM is working as expected. More specifically, we observe that univariate BD I TWAS hits that were identified as hits for the BD factor were far more significant for BD II than genes that were hits for BD I but not the overarching BD factor. Thus, T-SEM is functioning to identify patterns of gene expression specifically associated with shared risk pathways across the BD subtypes. No Q_Gene_ hits surpassed a Bonferroni corrected significance threshold. A list of the top 10 most significant Q_Gene_ hits is provided in Supplementary Table 5, many of which showed divergent directions of effects across subtypes.

**Fig. 3.**
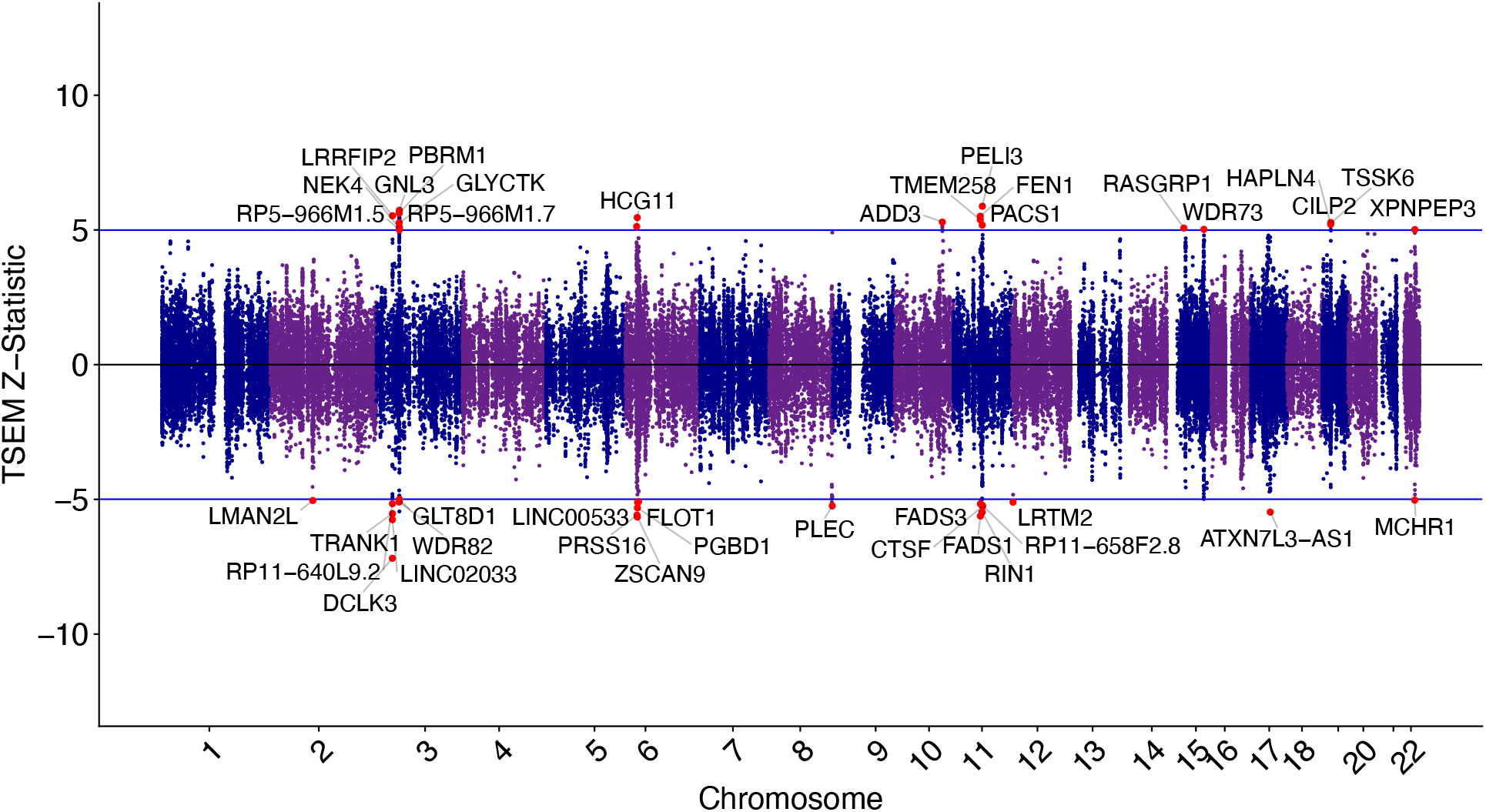
Miami Plot of Gene Expression Hits on the Bipolar Factor. The top and bottom blue bar represents *Z*-statistics surpassing a Bonferroni corrected significance threshold. Positive and negative values depict upward and downward patterns of gene expression associated with the BD factor, respectively. The most significant gene across tissue types are labeled and colored as red dots.

## Discussion

Providing support for a diagnostic manual that describes separate BD subtypes, the current findings identified 18 clinically relevant traits and disorders that had significantly discrepant genetic overlap with BD I and II. Challenging the notion that BD II is simply a less severe presentation along the BD continuum, BD II was found to have significantly larger genetic correlations with negative outcomes, with the sole exception of a larger genetic correlation between SCZ and BD I. Moreover, the BD II genetic correlation with SCZ was still sizeable. Although BD I’s greater association with SCZ may be interpreted as representing a more severe clinical outcome than the external traits more associated with BD II, it is striking that BD I’s elevated association with SCZ does not also result in greater downstream associations with the other outcomes examined.

The specific traits found to be more strongly associated with BD II can generally be summarized as reflecting different dimensions of the internalizing space, including the disorders themselves (i.e., anxiety disorders; MDD), their symptoms (insomnia; feeling fed up; feeling worried), and their clinical correlates (loneliness; negative self-ratings of health). These results are broadly consistent with prior phenotypic findings indicating larger associations, relative to BD I, between BD II and: frequency of depressive episodes (Baek et al., 2011), comorbid anxiety disorders (Mantere et al., 2006), general psychiatric comorbidity (Karanti et al., 2020), reduced social functioning (Baek et al., 2011), and insomnia (Steinan et al., 2016). Though it did not surpass Bonferroni corrected significance threshold, the point estimate for the genetic correlation with BD II was higher relative to BD I for both suicide attempts and self-harm, which we highlight as this has also been described phenotypically (Tondo et al., 2022). Given the pattern of current and prior findings, it is perhaps unsurprising that BD II has also been linked to greater burden of illness (Dell’Osso et al., 2015).

Theoretically informed follow-up analyses with MDD and SCZ as correlated predictors of each BD subtype indicate that the BD subtypes continue to exhibit unique contributions from these two disorders when modeled jointly. In addition, we find that BD subtypes are not simply blends of MDD and SCZ, with approximately half of the genetic variance unexplained by these two predictors. These large residual genetic variance components were also found to contain both shared and unique signal across the BD subtypes. Taken together, this tentatively supports bipolar disorder as a separate diagnostic class with distinguishable subtypes within this class. The considerable proportion of unique variability within each subtype supports their stratification in recent GWAS of the overarching disorder class. Further stratification in future GWAS of BD may also increase the power to detect additional divergences, whereas collapsing across the two disorders will mask subtype-specific signal and obscure relationships with external traits. Indeed, the most recent BD GWAS stratifying the disorder by subtype increased the SNP-based heritability estimate for BD I relative to the overarching disorder class (Mullins et al., 2021).

Epidemiological meta-analyses have estimated overall lifetime comorbidity rates of 17% across ADHD and BD (Schiweck et al., 2021). Although extant literature investigating comorbidity between BD and ADHD has stratified one of the two disorder classes by subtype, with ADHD distinguished by adult (aADHD) and childhood (cADHD) diagnosis, few studies have stratified both BD and ADHD simultaneously. When stratifying by BD subtypes, meta-analyses find no significant differences in ADHD comorbidity between BD I and II (Schiweck et al., 2021). Interestingly, our results are contrary to those found in epidemiological meta-analyses and indicate that genetic signal from BD II is likely driving the relationships previously observed between BD and the broader ADHD construct (Schiweck et al., 2021). In addition, our results support the notion that shared genetic liability is influencing comorbidity between these disorder classes. Indeed, genetic epidemiology has provided support for shared genetic risk factors, with relatives of BD probands exhibiting a heightened risk for ADHD (Faraone et al., 2012).

Results from T-SEM characterize patterns of gene expression that are specifically associated with the shared risk pathways across these two subtypes. Reaffirming the grouping of these subtypes within an overarching bipolar disorder class, we identify 41 unique gene IDs associated with general bipolar risk. At the same time, many genes were significant for BD I, but not the overarching BD factor, indicating again that BD is most appropriately modeled as two separate subtypes characterized by both shared and unique risk pathways. We consider here the prior research on a few of the top BD factor hits. The most significant hit, *DCLK3*, was also identified in the most recent BD GWAS and is a druggable target that codes for serine/threonine-protein kinase (Mullins et al., 2021; Wishart et al., 2018). Knockdown of gene expression for another top hit, *GNL3*, in human neuronal cell cultures was shown to cause aberrant proliferation and differentiation (Meng et al., 2020). Lastly, *PBRM1* has been previously found to be related to both BD and SCZ risk (Ripke, 2014; Stahl et al., 2019), with knockdown of expression of this gene in rat models resulting in a reduction of mushroom spine density in pyramidal neurons (Yang et al., 2020).

### Limitations and Future Directions

Several limitations of this work should be noted, with the most important being the restriction to only European samples based on limited data availability in other ancestries. To increase the representativeness of these findings we hope that, as datasets increase in size over the next several years, these analyses can be extended to other ancestral groups. An interpretive caveat of this work stems from recently described potential bias in genetic correlations due to cross-trait assortative mating (Border et al., 2022). However, due to the magnitude of genetic correlation and phenotypic similarity between these subtypes, cross-trait assortative mating alone is highly unlikely to account for the entirety of the genetic overlap (Grotzinger & Keller, 2022). Misdiagnosis represents another potential source of bias. For example, misdiagnosis of BD I as BD II (and vice versa) would inflate their genetic correlation (Wray et al., 2012), which in turn may underestimate the etiological divergences reported above. Conversely, misdiagnosis of MDD as BD II will decrease the genetic correlation between BD subtypes, which in turn may artificially inflate the observed divergences. As GWAS datasets increase in size, future work should seek to include schizoaffective disorder bipolar type in subsequent analyses of etiological divergences of disorders within the BD diagnostic class. Although we did not identify any significant Q_GENE_ hits, we were likely underpowered for these analyses and future work should continue to reevaluate patterns of gene expression that may underly genetic divergence across the BD subtypes. Finally, we highlight that the current conclusions should be interpreted as specifically reflecting common variant genetic architecture for SNPs with a MAF > 1%. While a recent whole exome sequencing study of bipolar disorder found a similar pattern of findings across subtypes, this should be reevaluated in future work as the results were noted by the study authors to be tentative given the relatively small subtype sample sizes (Palmer et al., 2022).

## Conclusions

We apply Genomic SEM to explore shared and unique genetic architecture across BD subtypes. At the level of genetic overlap of common SNPs, BD II was found to exhibit greater associations with phenotypes characteristic of internalizing disorders (i.e., anxiety, insomnia, and low physical activity), psychiatric symptoms, and adverse physical health outcomes. At the level of tissue-specific gene expression, T-SEM results uncovered 41 unique genes associated with signal shared across BD subtypes, with extant the literature implicating the top hits in key neuronal functions. Through a genetic lens, these results collectively support a diagnostic system that distinguishes these subtypes, while challenging the illness severity conception of BD that places BD II at the lower end of severity. The genetic divergence implicated by our results also strongly supports treating these as two separate entities in future genetic studies to avoid obscuring subtype specific risk pathways.

## Data Availability

The summary statistics for bipolar disorder I and II are available from the PGC website. The source for each external trait is listed in the supplementary tables.

https://pgc.unc.edu/for-researchers/download-results/

## Acknowledgments

The current analyses would not have been possible without the enormous efforts put forth by the investigators and participants from data resources such as the Psychiatric Genetics Consortium, iPSYCH, and UK Biobank.

## Financial Support

ADG and JL are supported by NIH Grant R01MH120219. ADG and IF are supported by NIA Grant RF1AG073593.

## Ethical Standards

All authors declare no competing financial interests or potential conflicts of interest.

## Notes

### Competing Interest Statement

The authors have declared no competing interest.

### Author Declarations

This study only used publicly available summary statistics that are available through the Psychiatric Genomics Consortium, Pan-UK Biobank, or a source provided by the original publication.

