## Supplemental Figure 1 for "Genomic SEM Applied to Explore Etiological Divergences in Bipolar Subtypes"

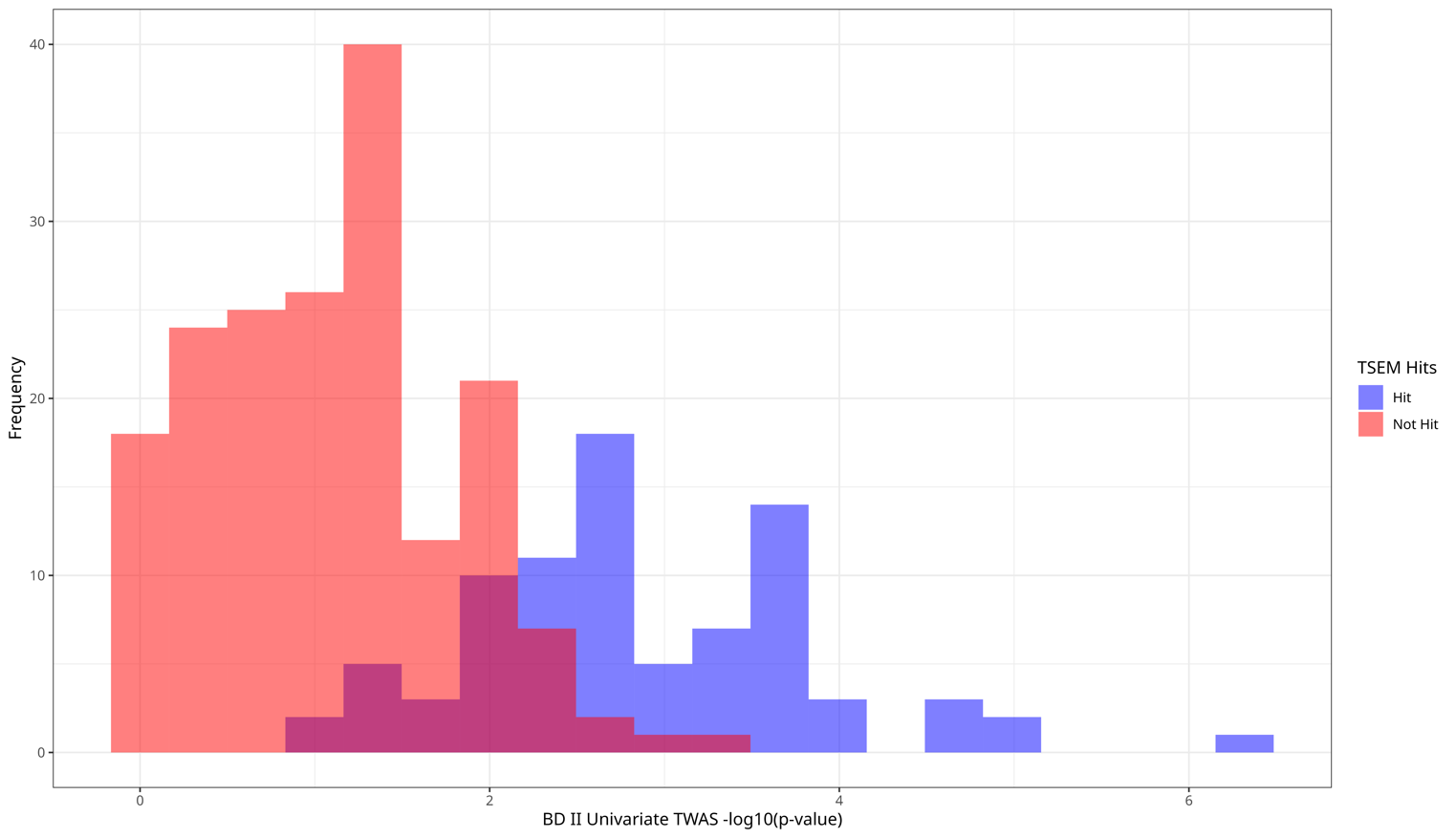
 **Supplementary Figure 1. BD II -log10(*p*-values) for BD I TWAS hits.** Blue bars represent BD II univariate TWAS *p*-values for BD I gene expression hits that are T-SEM gene expression hits. Red bars represent BD II univariate TWAS *p*-values for BD I gene expression hits not T-SEM gene expression hits.
